# Personal protective equipment for reducing the risk of COVID-19 infection among healthcare workers involved in emergency trauma surgery during the pandemic: a systematic review protocol

**DOI:** 10.1101/2020.09.26.20201954

**Authors:** Dylan P Griswold, Andres Gempeler, Angelos Kolias, Peter J. Hutchinson, Andres M. Rubiano

## Abstract

**Objective:** The objective of this broad evidence synthesis is to identify and summarize the available literature regarding the efficacy of different personal protective equipment (PPE) for reducing the risk of COVID-19 infection in health personnel caring for patients undergoing trauma surgery in low-resource environments (LREs).

**Introduction:** Many healthcare facilities in low-and middle-income countries are inadequately resourced and may lack optimal organization and governance, especially concerning surgical health systems. COVID-19 has the potential to decimate these already strained surgical healthcare services unless health systems take stringent measures to protect healthcare workers from viral exposure and ensure the continuity of specialized care for the patients.

**Inclusion criteria:** This review will preferentially consider systematic reviews of experimental and quasi-experimental studies, as well as individual studies of such designs, evaluating the effect of different PPE on the risk of COVID-19 infection in healthcare workers (HCWs) involved in emergency trauma surgery.

**Methods:** We will conduct several searches in the L·OVE (Living OVerview of Evidence) platform for COVID-19, a system that performs automated regular searches in PubMed, Embase, Cochrane Central Register of Controlled Trials (CENTRAL), and over thirty other sources. The search results will be presented according to the Preferred Reporting Items for Systematic Reviews and Meta-Analyses (PRISMA) flow diagram. Critical appraisal of the eligible studies for methodological quality will be conducted. Data will be extracted using the standardized data extraction tool in Covidence. Studies will, when possible, be pooled in a statistical meta-analysis using JBI SUMARI. The Grading of Recommendations, Assessment, Development, and Evaluation (GRADE) approach for grading the certainty of evidence will be followed, and a Summary of Findings (SoF) will be created.

**Systematic review registration number:** CRD42020198267

## INTRODUCTION

Many healthcare facilities in low-and middle-income countries are inadequately resourced. COVID-19 has the potential to decimate these already strained surgical healthcare services unless health systems take stringent measures to protect healthcare workers (HCWs) from viral exposure. A recent study showed that 15.6% of confirmed COVID-19 patients are symptomatic and that nearly half of patients with no symptoms at detection time will develop symptoms later.[1] Furthermore, the preoperative evaluation of emergency trauma patients is limited. These factors impede and confound diagnostic triage. Improper infection prevention may create a ‘super-spreader’ event in a high-volume healthcare facility or reduce available personnel. Consequently, the infection control strategy of trauma surgery staff is a top priority.

To take care of patients, providers must first take care of themselves. Personal protective equipment (PPE) is paramount to protect health care workers from contracting the virus and becoming disease carriers. Basic recommended PPE for trauma surgery staff of high-income country facilities include: 1) a surgical mask or better for all personnel interacting with patients and in the OR (including cleaning staff); 2) N95 or better mask for all staff in close contact with the patients (<6 feet away); 3) PAPR for aerosolising and high-risk procedures (ear, nose, throat, thoracic, and transsphenoidal neurosurgery operations); 4) universal testing of patients pre-operatively to enable appropriate PPE use, and 5) changing scrubs after every procedure.[2] These recommendations are suitable for high-resource settings but are less feasible in low-resource settings. A rapid-turnaround survey of 40 healthcare organisations across 15 LMICs revealed that 70% lack PPE and COVID-19 testing kits, and only 65% of the respondents showed confidence in hospital staff’s knowledge about precautions to be taken to prevent COVID-19 infection among hospital personnel.[3]

Some resource-adjusted recommendations include the use of cloth masks and bandanas. While innovative, their moisture retention, reusability, and filtration are considered very inferior to N95, and other masks.[4] What is most needed is evidence guided recommendations for PPE use and COVID-19 screening in LMICs surgical systems where resources are either limited or unavailable. HCWs have been instructed to consider refraining from caring for patients in the absence of adequate PPE availability.

A preliminary search of PROSPERO, MEDLINE, the Cochrane Database of Systematic Reviews, and the *JBI Database of Systematic Reviews and Implementation Reports* was conducted, and no current or underway systematic reviews on the topic were identified.

The primary objective of the review is to summarise the effects of different personal protective equipment in reducing the risk of COVID-19 infection of health personnel caring for patients undergoing trauma surgery. The purpose of the review is to inform recommendations for the rational use of PPE in emergency surgery staff, particularly in low resources environments where PPE shortages and high costs are expected to hamper the safety of HCWs and affect the care of trauma patients. A preliminary search of PROSPERO, MEDLINE, the Cochrane Database ofSystematic Reviews, and the JBI Database of Systematic Reviews and Implementation Reports was conducted, and no current or underway systematic reviews on the topic were identified.

## REVIEW QUESTIONS

What is the available evidence on the effects of various personal protective equipment (PPE) in reducing the risk of COVID-19 infection of health personnel caring for patients undergoing trauma surgery? We are also interested in data on the costs associated with the use of PPE since it is a vital aspect to consider when generating recommendations for low-resources environments.

## INCLUSION CRITERIA

### Participants

The review will preferentially include studies involving healthcare workers in emergency trauma surgery during the COVID-19 pandemic. Given the likelihood that reports on this specific population are scarce or even non-existent, if not available or insufficient we will consider studies of healthcare workers in any procedural and in-hospital setting such as ER and critical care management in COVID-19. Studies summarizing the available evidence for other viral respiratory illnesses will be considered if COVID-19 evidence is not available and the setting reported is trauma surgery.

### Intervention(s)

Different types of PPE used for treating patients requiring emergency trauma surgery.

### Comparator(s)

Comparators of interest are no PPE use and different types of PPE.

### Outcomes

This review will consider studies that include the following outcomes: Risk of contagion to health personnel involved in the care of the described population during the COVID- 19 pandemic; differences in surgical field vision; expressed as incidence differences, risk ratios or odds ratios.

If available, we will also extract and summarize information on cost associated with differenttypes of PPE. All outcomes will be summarized narratively

### Types of studies

This review will consider systematic reviews of experimental and observational studies, and experimental or observational studies if not included in systematic reviews that fulfilled population and intervention criteria. We will also include reports of costs associated with the use of PPE and reports on implementation strategies that could inform recommendations for low resource settings. Only studies published in English or Spanish will be included. We will include preprint studies identified in our search, but no ongoing studies will be considered.

## METHODS

A protocol of this review following the PRISMA statement was registered in the International Prospective Registry of Systematic Reviews (PROSPERO; CRD42020198267). This review was conducted following the JBI methodology for systematic reviews of aetiology and risk.[5]

### Search strategy

We will conduct several searches in the L·OVE (Living OVerview of Evidence) platform for COVID-19, a system that performs automated regular searches in PubMed, Embase, Cochrane Central Register of Controlled Trials (CENTRAL), and over thirty other sources. We will search for systematic reviews and randomized trials evaluating the effect of different personal protective equipment (PPE) on the risk of COVID-19 infection in personnel involved in emergency trauma surgery during the pandemic. Other in-hospital clinical settings will be considered for inclusion and synthesis if evidence for trauma surgery setting is not available. Different clinical settings will be treated as subgroups from which extrapolation will be possible when considered adequate. Non-randomized studies will be considered if systematic reviews and RCTs are not available, or if they report data on costs associated with the implementation of the interventions of interest. We will include preprint studies identified in our searches, but no ongoing studies will be considered. Ongoing studies will be counted as excluded studies in the corresponding tables and PRISMA diagram.

### Information sources

The databases to be searched include the L·OVE (Living OVerview of Evidence) platform for COVID-19, a system that performs automated regular searches in PubMed, Embase, Cochrane Central Register of Controlled Trials (CENTRAL), and over thirty other sources. When compared to manual searches, this platform consistently identifies all the available studies associated with the terms of interest. It allows for a fast (automated) search that is easy to update - a crucial element given the urgent need to answer the research question rapidly and thoroughly.

### Study selection

Following the search, all identified citations will be collated and uploaded into EndNoteX9 (Clarivate Analytics, PA, USA). The citations will then be imported into JBI SUMARI for the review process. Two independent reviewers will examine titles and abstracts for eligibility. The full text of selected studies will be retrieved and assessed. Full-text studies that do not meet the inclusion criteria will be excluded, and a list of such excluded studies will be provided. Disagreements between the reviewers during title and abstract screening or full-text screening, will be resolved by consensus, or with a third reviewer. The results of the search will be reported in full in the final report and presented in a Preferred Reporting Items for Systematic Reviews and Meta-analyses (PRISMA) flow diagram.[6]

### Assessment of methodological quality

Eligible studies will be critically appraised by one reviewer and verified with the other. We will use the AMSTAR tool to assess the risk of bias in systematic reviews, the Cochrane risk of bias tool (RoB2) for RCTs, and the ROBINS-I tool for non-randomized studies.[7–9] Risk of Bias will be assessed only for the primary outcome: infection of healthcare workers. The results of the risk of bias assessment will be reported narratively and inform grading of evidence summarized in SoF tables. Disagreements will be solved by consensus or by a third reviewer.

### Data extraction

Data will be extracted from the included studies by a reviewer and verified by a second reviewer using a data extraction tool from JBI SUMARI.[5] The data extracted will include specific details about the populations, study methods, interventions, and outcomes of significance to the review question and specific objectives. Disagreements will be solved by consensus.

### Data synthesis

Studies will be summarized narratively. Effect sizes from systematic reviews and from individual studies not included in them, will be expressed as odds ratios (for dichotomous data) with their 95% confidence intervals. Decision rules regarding data extraction for situations where data are:

- reported at multiple time points: include all and note time of measurement for each.
- multiple “doses”: N/A – (all types of PPE will be recorded)
- multiple exposures are compared (e.g., ever exposed, frequency of exposure): as mentionedbefore, exposure will relate to the clinical setting of healthcare workers studied and will be classified accordingly.

For summarizing results from other settings different to trauma surgery, effect of PPE will be summarized by subgroups according to different clinical settings.

### Assessing certainty in the findings

The Grading of Recommendations, Assessment, Development, and Evaluation (GRADE) approach for grading the certainty of evidence will be followed, and a Summary of Findings (SoF) will be created using online software GRADEPro GDT 2020 (McMaster University, ON, Canada).[10,11] The SoF will present the following information where appropriate: absolute risks for the treatment and control, estimates of relative risk, and quality of the evidence based on the risk of bias, directness, heterogeneity, precision, and risk of publication bias. The outcomes reported in the SoF will be risk of COVID-19 infection.

## Data Availability

All data will be made available upon request

## Acknowledgements

None to report

